# Protocol for the 2026 Nationwide Survey of Dementia Specialists on the Real-World Implementation of Anti-Amyloid Antibody Therapies in Japan: Clinical Practice, Blood Biomarkers, and Preference Experiments

**DOI:** 10.64898/2026.06.30.26356494

**Authors:** Kenichiro Sato, Yoshiki Niimi, Saki Nakashima, Ataru Igarashi, Atsushi Iwata, Kensaku Kasuga, Kiyotaka Nemoto, Shinji Higashi, Shuichi Awata, Manabu Ikeda, Takeshi Ikeuchi, Takeshi Iwatsubo, Tetsuaki Arai

**Author notes:** **Correspondence:** Yoshiki Niimi, M.D., Ph.D., Dementia Inclusion and Therapeutics, The University of Tokyo Hospital. 7-3-1 Hongo, Bunkyo-ku, Tokyo 113-8655, Japan.

## Abstract

**Background:** Anti-amyloid antibody therapies have changed the clinical pathway for early Alzheimer disease (AD). Lecanemab and donanemab are now clinically available in many countries, including Japan, and their use requires biomarker confirmation, repeated magnetic resonance imaging monitoring, management of amyloid-related imaging abnormalities, infusion capacity, staff resources, and shared decision-making. In Japan, these treatments are provided under the universal public health insurance system and are regulated by the optimal use guidelines. Therefore, it is important to understand not only the number of treated patients but also how specialists perceive clinical, logistical, and policy challenges in routine practice.

**Objective:** This paper describes the protocol for a nationwide anonymous online survey of dementia specialists in Japan. The survey aims to evaluate the real-world implementation of anti-amyloid antibody therapies, including current clinical practice, facility readiness, perceived barriers, possible policy solutions, and physician preferences assessed using a discrete choice experiment and best-worst scaling.

**Methods:** This is a prospective, cross-sectional, anonymous online survey using Google Forms. The survey targets board-certified specialists of the Japanese Society for Dementia Research and the Japanese Psychogeriatric Society, with a main focus on physicians who have completed the official training course required for anti-amyloid antibody therapy. The questionnaire includes items on respondent and facility characteristics, perceived value of treatment, treatment experience, diagnostic and eligibility assessment, amyloid and *APOE* testing, MRI monitoring, infusion capacity, continued-administration facilities, blood-based biomarkers, preclinical AD, a discrete choice experiment, and best-worst scaling. Among respondents routed to the DCE section, the discrete choice experiment asks respondents to choose between hypothetical anti-amyloid antibody treatment profiles for early AD, defined by expected efficacy, risk of amyloid-related imaging abnormalities requiring treatment interruption or discontinuation, treatment duration, visit frequency, waiting time, and monthly out-of-pocket cost. Best-worst scaling evaluates the relative importance of policy and system-level solutions.

**Results:** Data collection started on June 3, 2026, and is planned to close on June 30, 2026. This protocol was prepared before data lock and before any outcome analyses. The main results will be reported after data cleaning and analysis according to the prespecified analysis plan.

**Conclusions:** This protocol describes a nationwide survey designed to clarify clinical, logistical, and policy challenges in the implementation of anti-amyloid antibody therapies in Japan. By publishing the survey design and analysis plan before data lock, this study aims to improve transparency and interpretability. The findings will help identify where Japanese dementia specialists perceive bottlenecks in diagnosis, biomarker testing, safety monitoring, infusion delivery, continued administration, and reimbursement. They may also inform policy discussions on *APOE* testing, blood-based biomarkers, regional care coordination, and service reimbursement for anti-amyloid antibody therapy.

## Introduction

The approval and clinical use of lecanemab and donanemab have made biomarker-confirmed treatment pathways a routine consideration for early symptomatic Alzheimer disease (AD). Lecanemab showed a slowing of clinical decline in early AD with confirmed amyloid pathology in the Clarity AD trial, and donanemab also showed clinical benefit in early symptomatic AD in TRAILBLAZER-ALZ 2 [vanDyck2023; Sims2023]. These treatments have created a clinical pathway in which early diagnosis, biomarker confirmation, safety monitoring, and long-term treatment logistics are all required. Appropriate use recommendations for lecanemab and donanemab describe how these drugs should be used in routine practice, including patient eligibility, amyloid confirmation, *APOE* genotyping, MRI monitoring for amyloid-related imaging abnormalities, infusion procedures, and discussion of treatment benefits and risks [Cummings2023; Rabinovici2025]. This therapeutic pathway is also consistent with the recent shift toward biomarker-based diagnosis and staging of AD, in which biological evidence of AD pathology is increasingly central to clinical classification and treatment decision-making [Jack2024].

The implementation of anti-amyloid antibody therapies is complex because it depends on many components of the health care system. Patients need to be identified at an early symptomatic stage, undergo amyloid confirmation by positron emission tomography (PET) or cerebrospinal fluid (CSF) testing, receive repeated magnetic resonance imaging (MRI) scans, and have access to clinicians who can evaluate and manage ARIA. Health systems also need infusion capacity, trained staff, care coordination, reimbursement pathways, and continued-administration facilities. Previous modeling and readiness studies suggested that specialist capacity, biomarker testing, infusion delivery, and system coordination may become bottlenecks when disease-modifying therapies are introduced at scale [Mattke2022; Anderson2022; Walsh2024]. The possible implementation of blood-based biomarkers (BBMs), particularly plasma phosphorylated tau markers such as p-tau217, may reduce some diagnostic bottlenecks, but it also introduces new clinical, operational, and ethical questions related to assay performance, clinical context, disclosure, and health-system capacity [Mielke2024; Dyer2024; Palmqvist2025].

Japan is a relevant setting for evaluating implementation because lecanemab and donanemab are provided within a universal public insurance system and are governed by national optimal use guidelines. Lecanemab and donanemab are used under the national public health insurance system, and their use is guided by Japanese optimal use guidelines (OUGs) that specify patient-level, physician-level, facility-level, imaging, biomarker, and safety requirements [MHLW2023; MHLW2024]. Japan has a high density of MRI equipment and high MRI utilization compared with other countries [Aoyama2025]. However, safe implementation of anti-amyloid antibody therapy requires not only MRI availability but also standardized MRI protocols, ARIA interpretation expertise, and institutional MRI management systems [Kakeda2025]. A previous Japanese preparedness study also suggested that perceptions of anti-amyloid therapy implementation may differ between the public and specialists [Sato2024]. Therefore, Japan is a useful case for evaluating how disease-modifying AD therapies are implemented within a universal health insurance system.

A previous nationwide specialist survey in Japan described the first-year real-world adoption of lecanemab [Sato2025]. That study included 311 specialists who collectively treated 3259 patients with lecanemab and showed that early access appeared feasible, but that infrastructure and reimbursement barriers remained important. In particular, infusion space, staffing, continued-administration facilities, reimbursement for infusion-related services, and insurance coverage for *APOE* testing were identified as important issues. Since that earlier survey, the clinical and policy environment has changed. Donanemab has been introduced, blood-based biomarkers are moving toward clinical implementation, *APOE* testing remains an important safety and counseling issue, subcutaneous maintenance treatment is being discussed, and preclinical AD is increasingly being discussed as a potential target for future prevention or early-intervention strategies [Rabinovici2025; Mielke2024; Mattke2020; Sperling2011; Ressa2026].

Standard survey questions are useful for describing frequencies and opinions, but they may not fully capture trade-offs. For example, a physician may accept a higher ARIA risk if the expected benefit is larger, or may prefer a treatment with a smaller benefit if the visit burden and out-of-pocket cost are lower. Similarly, many policy solutions may be considered important, but ordinary multiple-choice questions do not easily show their relative priority. Discrete choice experiments (DCEs) are widely used in health care to quantify trade-offs between attributes of treatments, services, or policies [Lancsar2008; Bridges2011]. Best-worst scaling (BWS) can also be used to evaluate the relative importance of items by asking respondents to choose the most and least important options from repeated sets [Flynn2007; Flynn2010].

For these reasons, the present survey includes both a DCE and a BWS component. In the DCE, respondents who reach the DCE section are asked to choose between hypothetical anti-amyloid antibody treatment profiles for early AD. The profiles differ by expected efficacy, risk of ARIA requiring treatment interruption or discontinuation, treatment duration, visit frequency, waiting time until treatment initiation, and monthly out-of-pocket cost. Similar DCE approaches have been used to evaluate Alzheimer treatment preferences among caregivers and neurologists, including attributes related to clinical effects, ARIA, treatment duration, and administration route or frequency [Dranitsaris2023]. The design, analysis, and reporting of DCEs in health care require careful consideration of experimental design, sample size, model specification, and transparent reporting [Johnson2013; deBekkerGrob2015; Hauber2016; Ride2024]. In the BWS component, specialists are asked to prioritize policy and system-level solutions, including drug development, administration route improvement, medical and care professional education, shared decision-making reimbursement, infusion-related reimbursement, *APOE* testing coverage, blood-based biomarker implementation, regional care coordination, and non-pharmacological intervention research.

Publishing the protocol before data lock and outcome analysis can improve transparency and help readers interpret the final results. Web-based surveys need clear reporting of recruitment, consent, sampling, prevention of duplicate responses, questionnaire structure, mandatory items, response rates, and data cleaning procedures [Eysenbach2004]. The Checklist for Reporting of Survey Studies also emphasizes transparent reporting of survey design, participants, recruitment, ethical considerations, data handling, and analysis [Sharma2021]. Although the present study is not a clinical trial, the broader principle that a study protocol should describe the rationale, methods, organization, and ethical considerations before analysis is also consistent with established protocol guidance [Chan2013].

The objective of this paper is to describe the protocol for a nationwide anonymous online survey of dementia specialists in Japan on the real-world implementation of anti-amyloid antibody therapies. The protocol describes the study design, participants, recruitment, questionnaire structure, DCE and BWS components, outcomes, data management, statistical analysis plan, ethical considerations, and dissemination plan.

## Methods

### Study Design

This study is a prospective, cross-sectional, anonymous online survey of dementia specialists in Japan. It is designed as a survey protocol paper and does not report outcome analyses. The survey was developed to evaluate real-world clinical practice and implementation challenges related to anti-amyloid antibody therapies, including lecanemab and donanemab. The survey is being conducted using Google Forms. Data collection started on June 3, 2026, and is planned to close on June 30, 2026. Reminder emails will be sent during the survey period to improve participation. This protocol was prepared before data lock and before any analysis of survey outcomes.

The study is conducted as part of a Ministry of Health, Labour and Welfare research project on social issues associated with advances in dementia medicine (https://mhlw-grants.niph.go.jp/project/176226) (PI: Tetsuaki Arai). The survey is implemented with cooperation from the Japanese Society for Dementia Research (https://dementia-japan.org) (president: Takeshi Ikeuchi) and the Japanese Psychogeriatric Society (https://www.rounen.org) (president: Manabu Ikeda).

### Participants and Recruitment

The target population is physicians who are board-certified specialists of the Japanese Society for Dementia Research or the Japanese Psychogeriatric Society. The main target group is specialists who have completed the official training course required for the administration of anti-amyloid antibody therapy for AD in Japan. However, physicians who have not completed the training course, and physicians who have completed the course but do not currently administer anti-amyloid antibody therapies, are also allowed to respond to relevant sections of the survey.

The survey URL is distributed by email through the collaborating societies. Participation is voluntary. The invitation explains that the survey is anonymous, that email addresses are not collected, and that respondents are asked to avoid duplicate responses. Because the survey is anonymous, no direct personal identifiers, such as name, affiliation, or email address, are collected.

This survey is exploratory and descriptive. The target number of responses is approximately 200 to 300, based on feasibility, expected response patterns, and previous related surveys. This number is not based on a single confirmatory hypothesis test. For the DCE component, we used the commonly applied rule of thumb N > 500c/(t × a), where c is the largest number of levels for any attribute, t is the number of core choice tasks per respondent, and a is the number of alternatives per task [deBekkerGrob2015]. In this survey, c=3, t=10, and a=2, giving a minimum target of more than 75 respondents reaching the DCE core tasks. Mixed logit models and subgroup analyses will be interpreted as exploratory and will be conducted only if the achieved DCE sample size is sufficient. The final number of responses will depend on the actual response rate during the survey period.

### Survey Instrument

The questionnaire was developed by the research team based on previous work on anti-amyloid antibody therapy implementation [Sato2025], the Japanese optimal use guidelines, clinical experience, and policy issues identified in the first year after lecanemab launch [MHLW2023; MHLW2024]. The survey includes both standard survey questions and stated-preference components.

The questionnaire begins with an explanation of the study and an online consent item. Respondents who do not agree to participate are directed to the end of the survey. Respondents who agree proceed to the questionnaire. The survey then collects background information, including specialist qualifications, years of specialist experience, facility type, dementia medical center status, outpatient setting, prefecture, amyloid PET availability, and CSF testing availability.

The next section evaluates the perceived value of treatment with lecanemab or donanemab. Respondents are asked to rate the value of treatment from multiple perspectives, including medical value, care-related value, maintenance of daily life, response to patient and family preferences, hospital management, allocation of health care resources, and social health economic value. A BWS item is also used to compare the relative importance of these dimensions.

The survey then asks about treatment experience. Respondents are asked whether they currently use or have previously used lecanemab, donanemab, or both. According to their answers, they are routed to questions on facility type, including whether their main facility can provide initial treatment, only continued administration, no administration, or treatment outside public insurance.

For physicians with relevant treatment experience, the questionnaire asks about barriers to introduction, continued-administration facilities, reasons for refusal by continued-administration facilities, drug price as a barrier, waiting time from first consultation to treatment initiation, appointment waiting time, change in referrals or visits after drug introduction, and change in the number of patients being considered for treatment.

The survey also includes detailed questions on eligibility assessment and biomarker testing. These questions cover amyloid confirmation by PET or CSF, disclosure of amyloid test results, patient and caregiver understanding and acceptance of amyloid positivity, need for disclosure protocols, amyloid positivity rate in different patient groups, repeated amyloid testing, MRI field strength, MRI sequences for hemorrhagic changes, *APOE* testing, disclosure of *APOE* results, genetic counseling, and whether *APOE* results influenced treatment decisions.

The treatment delivery section asks about first administration in inpatient settings, outpatient infusion space, outpatient staff, continued-administration facility availability, treatment capacity compared with expected demand, expected impact of subcutaneous maintenance therapy, linkage with dementia medical centers, patient and caregiver satisfaction with medical cost and treatment effect, and use of financial support systems.

Additional sections cover safety-related practice, blood-based biomarkers, and preclinical AD. The blood-based biomarker section asks about the expected role of blood-based biomarkers as prescreening tools or as possible substitutes for PET or CSF testing. The preclinical AD section asks about attitudes toward the significance of possible treatment or preventive intervention in preclinical Alzheimer disease from medical, care-related, patient/family, insurance, and societal perspectives.

Most non-branching and non-consent questions are optional. This design was chosen because the survey is long and includes several specialized sections. Mandatory items are limited mainly to consent and key branching questions required to direct respondents to appropriate sections.

### Discrete Choice Experiment

The discrete choice experiment was designed to evaluate how dementia specialists trade off key attributes of anti-amyloid antibody therapy for early Alzheimer disease. Respondents who reach the DCE section are asked to imagine an amyloid-positive patient with mild cognitive impairment or mild dementia due to Alzheimer disease and to choose between two hypothetical treatments, treatment A and treatment B. The treatment profiles differed by six attributes: expected efficacy, risk of ARIA requiring treatment interruption or discontinuation, treatment duration, outpatient visit frequency, waiting time until treatment initiation, and monthly out-of-pocket cost. The DCE design was informed by published good-practice recommendations for constructing experimental designs, considering sample size, analyzing choice data, and reporting health-related DCEs [Johnson2013; deBekkerGrob2015; Hauber2016; Ride2024]. The efficacy levels were intended to create sufficient variation for evaluating trade-offs in hypothetical treatment profiles and should not be interpreted as direct estimates of the clinical effects of currently available drugs.

The DCE design was generated in two steps. First, a profile-level D-optimal design was used to select treatment profiles from the full factorial candidate set. The candidate set included six attributes with three candidate levels each, resulting in 729 (=3^6) possible profiles. Expected efficacy and outpatient visit frequency were treated as categorical attributes. ARIA risk, treatment duration, waiting time until treatment initiation, and monthly out-of-pocket cost were treated as scaled continuous attributes with linear effects in the design model.

From the 729 candidate profiles, 80 profiles were selected using the *optFederov* function in the R package *AlgDesign*, with 200 repeated searches and a fixed random seed. The calculated D-efficiency index of 0.783 refers to this profile-level selection under the prespecified design model before pairing into choice sets. Because the four numerical attributes were modeled linearly, the final selected profiles retained only the lower and upper candidate levels for these attributes. Therefore, in the implemented DCE tasks, ARIA risk appeared at 1% and 15%, treatment duration at 6 months and 3 years, waiting time until treatment initiation at 1 month and 6 months, and monthly out-of-pocket cost at JPY 10,000 and JPY 50,000. Expected efficacy and outpatient visit frequency retained all three candidate levels.

Second, the 80 selected profiles were paired into 40 two-alternative core choice sets. Pairing was performed as a constrained pairing procedure to avoid unintended dominance under the prespecified preference direction, so that neither alternative was clearly better than the other across all attributes in the core choice tasks. The 40 core choice sets were divided into 4 blocks of 10 core tasks, with the aim of maintaining approximate balance across attributes within blocks. The implemented design was therefore a pragmatic profile-level D-optimal design followed by constrained non-dominant pairing, rather than a prior-based choice-set-level D-optimal design for a multinomial logit information matrix. This pragmatic approach was selected because reliable prior preference estimates for Japanese dementia specialists were not available before the survey.

Each block was supplemented with 1 position-swapped duplicate task and 1 dominance task, resulting in 12 DCE tasks per respondent. The position-swapped duplicate task repeated one core choice set with the A/B labels reversed. This task was included to assess whether respondents chose the same treatment profile when the alternatives were presented in reversed order. Consistency will therefore be evaluated by whether the same treatment profile was selected, not by whether the same A or B label was selected. The dominance task was included to evaluate whether respondents selected the clearly preferable alternative under the prespecified preference direction. The dominance task will be used as a data-quality indicator rather than as an automatic exclusion criterion in the main analysis.

The assignment to DCE blocks was implemented in Google Forms using a pseudo-random allocation based on the parity of the respondent’s birth month and birth day. This method was selected because Google Forms does not provide built-in randomization functions suitable for complex block allocation. The birth month and birth day were asked only as even/odd categories and were not collected as full dates.

### Best-Worst Scaling

The best-worst scaling component was designed to evaluate the relative priority of possible policy and system-level solutions for the implementation of anti-amyloid antibody therapies. The candidate items were selected mainly from issues identified in our previous nationwide specialist survey conducted 1 year after lecanemab launch in Japan, which highlighted infrastructure and reimbursement barriers such as infusion space, staff resources, continued-administration facilities, *APOE* testing coverage, and additional reimbursement for infusion-related services [Sato2025]. In the present survey, we used best-worst scaling to move beyond whether each issue was considered important and to quantify the relative priority of these possible solutions.

The BWS item list includes 15 candidate solutions: promotion of new drug development, improvement of administration route and method, education for medical professionals, education for care professionals, reimbursement for shared decision-making when introducing anti-amyloid antibody therapy, outpatient chemotherapy-administration-style reimbursement add-on when administering anti-amyloid antibody therapy, reimbursement for home self-injection management if self-injection becomes available, reimbursement for continued-administration coordination, revision of optimal use guidelines, relaxation of testing restrictions, insurance coverage for *APOE* testing, implementation of blood-based biomarkers with relatively established evidence such as p-tau217, implementation of future blood-based biomarkers as evidence accumulates, development of regional care networks adapted to the anti-amyloid antibody era, and promotion of non-pharmacological intervention research.

The BWS design consists of 15 items, 4 groups, 6 tasks per group, and 5 items per task. The 4 groups represent alternative presentation versions, not item subsets. Within each group, all 15 items appear exactly twice across the 6 tasks, and each task contains 5 items. Therefore, each respondent assigned to the BWS component evaluates all 15 candidate solutions, although the task composition and item order differ by group. In each task, respondents are asked to choose the most important and least important item. This design reduces respondent burden while allowing comparison of the relative priority of policy solutions.

Assignment to BWS groups was implemented using the same practical Google Forms allocation variables as the DCE block assignment. Therefore, BWS group membership and DCE block assignment are treated as presentation-design factors, not as independent respondent subgroups. We do not plan to interpret differences between BWS groups or DCE blocks as substantive subgroup effects.

### Outcomes

The primary outcomes are current use of lecanemab and donanemab, facility readiness, perceived implementation barriers, diagnostic and treatment workflows, and support for policy solutions. Facility readiness includes amyloid testing, CSF testing, MRI monitoring, ARIA response capacity, infusion space, staff resources, and continued-administration facilities.

Secondary outcomes include physician preferences estimated from the DCE, relative importance of policy solutions estimated from BWS, attitudes toward *APOE* testing, attitudes toward blood-based biomarkers, and attitudes toward possible preventive treatment or intervention in hypothetical preclinical AD.

Feasibility outcomes include the total number of responses, completion rate, item-level missingness, branching-related nonresponse, DCE response rate, BWS response rate, dominance task pass rate, position-swapped duplicate task consistency, and response to the instructed-response attention-check item. These feasibility and data-quality outcomes will be used to evaluate whether the instrument was practical for a nationwide specialist survey.

### Data Management

Survey responses will be exported from Google Forms as a spreadsheet or CSV file. The research team will assign an analysis ID to each response. Direct personal identifiers are not collected. Variables will be processed according to a prespecified codebook.

Because the survey URL is distributed through collaborating academic societies, the exact number of physicians who opened the invitation email or viewed the survey page cannot be determined by the research team. Therefore, a view rate will not be calculated. If the number of invited society members or eligible specialists is available from the collaborating societies, we will report a response rate using that number as the denominator. Completion rate will be defined as the proportion of consenting respondents who reach the final survey page, and section-specific completion will be reported for major optional sections, including the DCE and BWS components.

Missing data will be classified into at least three categories: structurally missing because of branching, item nonresponse because the question was optional, and invalid or uninterpretable responses. Free-text responses will be reviewed and coded where appropriate. Numerical responses will be checked for implausible values. For multiple-choice questions, each selected option will be coded as a binary variable when needed.

For data-quality assessment, the survey includes one instructed-response attention-check item in the blood-based biomarker section. This item is coded as 069_000 in Table S1 and corresponds to Google Forms question 188 on page 73 of the exported survey form. It is placed immediately after the item on testing cost and before subsequent items on assay interpretation. The item states: “This is an attention-check item. For this question, please select ‘not very important.’” Respondents who do not select the instructed response will be flagged for data-quality assessment. Because this instructed-response item is placed after the DCE and BWS sections, it will be used as a general survey-level attention indicator and will not be interpreted as a direct attention check for the DCE or BWS tasks.

For DCE data, responses will be reshaped into a long format suitable for choice modeling. Each alternative in each choice task will be represented as one row, with variables for respondent ID, block, task ID, alternative label, selected alternative, task type, and attribute levels. Core tasks, position-swapped duplicate tasks, and dominance tasks will be identified separately.

The full questionnaire, codebook, DCE design matrix, and BWS design matrix are provided as supplementary files.

### Statistical Analysis Plan

Categorical variables will be summarized as counts and percentages. Continuous or count variables will be summarized using median and interquartile range, and mean and standard deviation when appropriate. For each question, the denominator will be reported clearly. Unless otherwise specified, percentages will be calculated using the number of valid responses to that question as the denominator.

Analyses will first describe the overall respondent population. Then, analyses will be stratified by treatment experience, facility type, dementia medical center status, initial-treatment facility status, continued-administration facility status, and availability of amyloid PET or CSF testing. Differences between groups will be evaluated descriptively. When appropriate, exploratory regression models will be used to evaluate associations between respondent or facility characteristics and selected outcomes.

For multiple-choice questions on implementation barriers and policy solutions, each option will be coded as selected or not selected. Logistic regression models may be used to estimate the association between treatment experience and selection of specific barriers or solutions. Results will be reported as odds ratios with 95% confidence intervals. Because this study is exploratory, interpretation will focus on effect size and confidence intervals rather than only P values.

The DCE will be analyzed using conditional logit models as the primary approach. The primary DCE analysis will use the 10 core choice tasks in each block. The position-swapped duplicate task and the dominance task will be used for data-quality assessment and sensitivity analyses, not for the main preference model. Mixed logit models may be used as a secondary approach if the number of responses is sufficient [Hauber2016]. If the number of respondents reaching the DCE section is below the rule-of-thumb threshold described above, the DCE results will be reported descriptively or interpreted with particular caution, and mixed logit and subgroup analyses will not be emphasized.

Expected efficacy and outpatient visit frequency will be modeled as categorical attributes. ARIA risk, treatment duration, waiting time until treatment initiation, and monthly out-of-pocket cost will be modeled as continuous linear attributes, consistent with the design model. Because treatment A and treatment B are unlabeled hypothetical alternatives, no alternative-specific constant will be included in the primary model. Attribute coefficients, direction of preference, and relative importance will be estimated. For expected efficacy, the ordering of estimated coefficients across levels will be examined descriptively to assess whether preferences are consistent with the assumed favorable direction. When appropriate, exploratory subgroup analyses may be conducted by treatment experience, facility type, or other respondent characteristics.

For DCE data-quality assessment, the dominance task pass rate will be defined as the proportion of respondents who selected the alternative that was preferable under the prespecified preference direction. Duplicate-task consistency will be defined as the proportion of respondents who selected the same treatment profile in the original and position-swapped duplicate tasks. Sensitivity analyses may be performed after excluding respondents who fail the dominance task, show inconsistent responses in the position-swapped duplicate task, or fail the instructed-response attention-check item.

The BWS data will first be summarized using count-based best-minus-worst scores. Conditional logit models may then be used to estimate the relative importance of policy and system-level solutions. Results will be shown as rankings and, where possible, scaled preference scores.

Missing data will not be imputed for the main descriptive analyses. For regression analyses, complete-case analysis will be used for the variables included in each model. Sensitivity analyses may be performed if item-level missingness is substantial.

### Ethics

The study has been approved by the Ethics Committee of the University of Tokyo Graduate School of Medicine (2024397NI-(1)). Online informed consent is obtained at the beginning of the survey. Respondents who do not agree to participate are directed to the end of the survey and are not asked to complete the questionnaire.

The survey is anonymous. Names, affiliations, and email addresses are not collected. Because the survey does not collect information that allows the research team to identify individual respondents, withdrawal of consent after survey completion is not possible. This is explained in the survey information page.

### Dissemination

The results will be disseminated through peer-reviewed publications, conference presentations, research reports, and feedback to relevant academic societies and policy stakeholders. The results may also be used to inform discussions on clinical implementation, reimbursement, *APOE* testing, BBMs, regional care coordination, and future research on anti-amyloid antibody therapies.

## Results

Data collection started on June 3, 2026, and is planned to close on June 30, 2026. Reminder emails are planned during the survey period. At the time of preparation of this protocol, data collection was ongoing, and data lock had not been performed.

No outcome analyses had been conducted at the time this protocol was prepared. All analyses will be performed after the survey period has ended, after data cleaning, and according to the analysis plan described above.

The final study results will be reported in several domains: respondent and facility characteristics, current use of lecanemab and donanemab, facility readiness and implementation barriers, diagnostic and eligibility assessment, amyloid and *APOE* testing, MRI and ARIA-related practice, infusion and continued-administration capacity, blood-based biomarker attitudes, preclinical AD attitudes, DCE results, and BWS results.

## Discussion

This protocol describes a nationwide anonymous online survey designed to evaluate the implementation of anti-amyloid antibody therapies among dementia specialists in Japan. The survey is intended to capture real-world clinical practice, facility readiness, perceived barriers, policy needs, and physician preferences in the rapidly changing AD disease-modifying therapy era.

The main value of this study is that it addresses several levels of implementation at the same time. Anti-amyloid antibody therapy is not only a pharmacological intervention. It also requires early identification of eligible patients, biomarker confirmation, repeated MRI monitoring, ARIA management, infusion delivery, staff resources, financial support, and coordination between initial-treatment and continued-administration facilities. These requirements can create bottlenecks at different points in the care pathway [Mattke2022; Anderson2022; Walsh2024]. By surveying specialists who are directly involved in dementia care, this study may clarify which barriers are most visible in routine practice.

Another important feature is that the survey includes both conventional questionnaire items and stated-preference components. Standard questions can describe how many respondents report a given problem or support a given solution. However, they cannot fully show how physicians trade off benefit, safety, treatment burden, access, and cost. The DCE is designed to address this limitation by asking respondents to choose between hypothetical treatment profiles. The BWS component is designed to show the relative priority of policy solutions, instead of only asking whether each solution is important. These methods are exploratory in this protocol, but they may provide information that is difficult to obtain from ordinary survey questions alone [Lancsar2008; Bridges2011; Flynn2007; Dranitsaris2023].

The present survey also extends the previous Japanese specialist survey conducted after the first year of lecanemab use. That earlier study suggested that access to lecanemab was feasible in many settings, but that infusion space, staff resources, reimbursement, *APOE* testing, and continued-administration facilities remained important barriers [Sato2025]. The current survey was designed for a later phase, when donanemab has become clinically relevant and when additional topics, such as blood-based biomarkers, subcutaneous maintenance treatment, and preclinical AD, are increasingly important.

The findings are expected to be directly relevant to reimbursement and service-design discussions in Japan. The survey specifically examines physician support for *APOE* testing coverage, blood-based biomarker implementation, reimbursement for shared decision-making and infusion management, coordination fees for continued-administration facilities, and development of regional care networks. Similar affordability concerns have been raised in broader discussions of anti-amyloid antibody therapy, suggesting that access, reimbursement, and public financing are central issues for sustainable implementation [Jonsson2023]. These issues are directly related to the sustainability of AD disease-modifying therapy within the Japanese public health insurance system. Findings may also help academic societies and policy stakeholders understand where further guidance, education, or system support is needed.

Clinically, the survey focuses on points at which treatment decisions may fail in routine practice: biomarker disclosure, *APOE* counseling, interpretation of modest treatment benefits, management of treatment burden, and communication about cost. Anti-amyloid antibody therapy requires careful communication with patients and caregivers because expected benefits are modest, treatment burden is substantial, and safety monitoring is necessary. The survey evaluates not only facility-level barriers but also how specialists perceive patient and caregiver understanding, acceptance of amyloid positivity, *APOE* disclosure, treatment cost, and treatment effect. These results may help improve shared decision-making and clinical communication in routine dementia care, particularly because biomarker disclosure and dementia-related treatment decisions often involve patients, caregivers, and clinicians with different informational needs and preferences [Daly2018; Wake2020; Liu2025].

This study has several strengths. It is nationwide, targets dementia specialists, includes both users and non-users of anti-amyloid antibody therapies, and uses an anonymous online format. The questionnaire covers a broad range of implementation domains, including clinical practice, facility capacity, biomarker testing, treatment delivery, continued administration, policy solutions, and future technologies. The study also includes a DCE and BWS module, which may provide additional information on trade-offs and relative priorities. The publication of this protocol before data lock and outcome analysis is intended to improve transparency.

However, several limitations should be considered. First, this is a voluntary online survey. Respondents who have a strong interest in anti-amyloid antibody therapy, or who have direct treatment experience, may be more likely to respond. Therefore, the results may not represent all dementia specialists in Japan. Second, because the survey URL is distributed through collaborating academic societies, the exact number of physicians who viewed the invitation or opened the survey page cannot be determined by the research team. Response rates and completion rates will therefore need to be interpreted according to the available denominator information. Third, the questionnaire is long and includes complex branching. Although this design allows detailed assessment of many domains, it may increase item nonresponse and reduce completion of optional sections such as the DCE and BWS. Fourth, because the survey is anonymous, duplicate responses cannot be completely excluded, although respondents are asked to avoid duplicate participation. Fifth, this is a cross-sectional survey. Associations between facility characteristics and perceived barriers should be interpreted as exploratory and not causal. Sixth, the DCE and BWS tasks use hypothetical scenarios. They can help estimate preferences and priorities, but they may not fully predict real clinical behavior or policy decisions. Seventh, the pseudo-random allocation based on even/odd birth month and birth day is a practical solution for Google Forms, but it is not equivalent to computer-generated randomization.

Several limitations specific to the DCE should also be considered. First, although the DCE candidate set included three candidate levels for all six attributes, four numerical attributes appeared only at their lower and upper levels in the final implemented tasks because these attributes were modeled as linear continuous variables in the design model. Therefore, the DCE will estimate linear preferences over the prespecified ranges for ARIA risk, treatment duration, waiting time, and monthly out-of-pocket cost. It will not evaluate nonlinear or threshold effects at the candidate middle levels, such as 5% ARIA risk, 1-year treatment duration, 3-month waiting time, or JPY 30,000 monthly out-of-pocket cost. Second, the upper efficacy level used in the DCE may exceed the effect sizes observed in current clinical trials of anti-amyloid antibody therapies. This level was included to create sufficient variation for preference estimation, but it may affect anchoring and external validity. Third, the interpretation of treatment duration may not be uniform across respondents. The design assumed that a shorter planned treatment duration was less burdensome, but some clinicians may interpret a longer duration as indicating longer continuation of benefit or treatment opportunity. Therefore, the coefficient for treatment duration will be interpreted cautiously as a stated preference for the presented treatment profile, rather than as a pure measure of treatment burden. Fourth, the DCE was shown only to respondents who were routed to the DCE section after treatment-experience and facility-status branching. Therefore, DCE results should be interpreted as preferences among this respondent subgroup, not necessarily among all consenting respondents. In addition, the DCE attribute for ARIA risk combined ARIA events requiring treatment interruption or discontinuation and did not distinguish ARIA-E from ARIA-H. Therefore, the DCE cannot estimate preferences for specific ARIA subtypes.

Despite these limitations, the survey can provide a structured snapshot of how Japanese dementia specialists are adapting to the combined introduction of lecanemab, donanemab, biomarker-based eligibility assessment, and facility-level safety monitoring. The final analyses will describe current practice and perceived barriers, compare responses across facility and treatment-experience groups, evaluate DCE-based treatment preferences, and rank policy solutions using BWS. The findings may also guide future studies that compare changes over time as AD disease-modifying therapy becomes more widely implemented.

Future surveys should also examine several topics that could not be fully addressed in the present questionnaire. First, *APOE* testing is likely to become a more important issue if insurance coverage or wider clinical implementation becomes feasible in Japan. More detailed information will be needed on how *APOE* testing is currently performed, how results are disclosed, how genetic counseling is provided, and how physicians judge the clinical value of *APOE* information in treatment decision-making. Second, the possible introduction of subcutaneous lecanemab maintenance therapy may change the clinical meaning of treatment burden, outpatient capacity, staff workload, and patient preference. Future surveys should evaluate how specialists expect subcutaneous treatment to affect treatment continuation, facility capacity, and regional care coordination. Third, the role of evidence-based non-pharmacological interventions should be evaluated in more detail, especially for patients who are not eligible for anti-amyloid antibody therapy or who decide not to receive such treatment, because cognitive stimulation approaches have shown benefits for cognitive function and quality of life in people with dementia [Spector2003; Woods2023]. These topics may become increasingly important as disease-modifying therapies are implemented more widely and as dementia care needs to integrate pharmacological treatment, diagnostic counseling, and long-term supportive care.

In conclusion, this protocol describes a nationwide anonymous online survey of dementia specialists in Japan on the real-world implementation of anti-amyloid antibody therapies. The survey integrates standard questionnaire items with a DCE and BWS to evaluate both implementation barriers and physician preferences. By clarifying the study design and analysis plan before data lock, this protocol aims to support transparent interpretation of the final results and to provide a foundation for future clinical, policy, and preference research in the AD disease-modifying therapy era.

**Figure 1.**
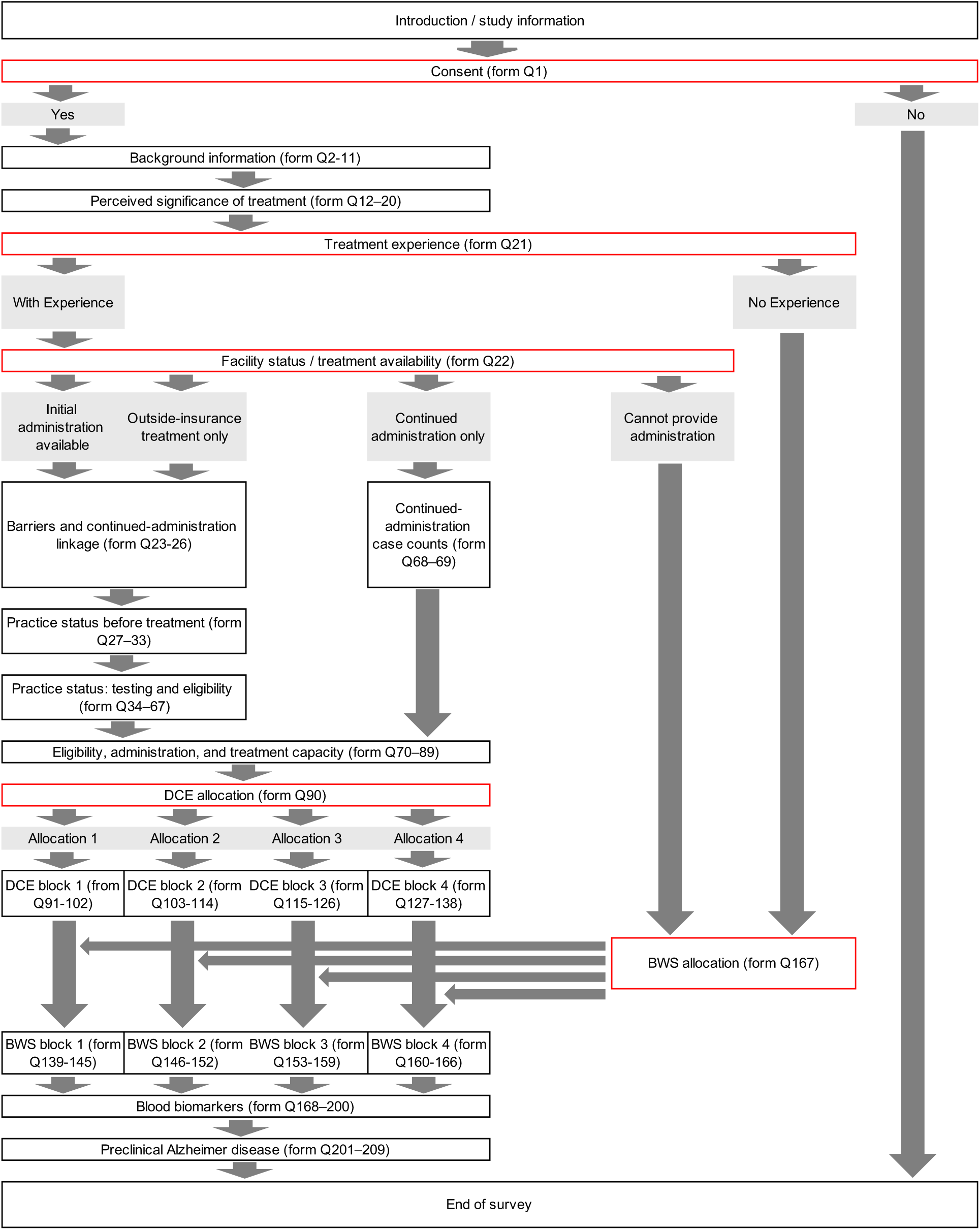
Overall survey flow and branching structure. This figure shows the planned flow of the anonymous online survey. After reading the study information, respondents provide online consent. Respondents who do not agree are directed to the end of the survey. Respondents who agree proceed to background questions and then to subsequent sections according to their training status, treatment experience, and facility type. Physicians routed to treatment-practice sections answer detailed questions on eligibility assessment, amyloid testing, *APOE* testing, MRI monitoring, infusion capacity, continued-administration facilities, safety-related practice, blood-based biomarkers, and preclinical Alzheimer disease. The discrete choice experiment and best-worst scaling components are presented near the end of the survey. **Abbreviations**: AD, Alzheimer disease; *APOE*, apolipoprotein E; BBM, blood-based biomarker; BWS, best-worst scaling; DCE, discrete choice experiment; MRI, magnetic resonance imaging.

**Figure 2.**
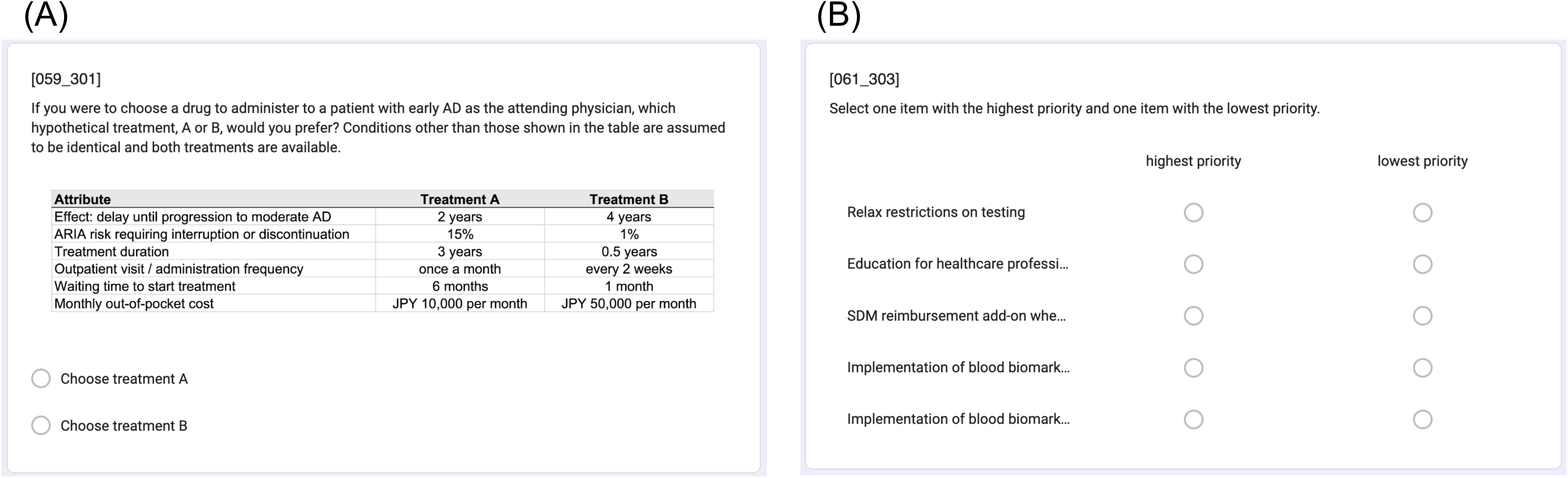
English-rendered examples of the discrete choice experiment and best-worst scaling components. (A) Example discrete choice experiment task. Respondents were asked to choose between two hypothetical anti-amyloid antibody treatment profiles for an early AD patient. (B) Example best-worst scaling task. Respondents were asked to select the highest- and lowest-priority items from a set of possible policy and system-level solutions. The actual survey was administered in Japanese; these panels are English-rendered reproductions prepared for readers. The wording is based on the English translations provided in the supplementary tables, and the task structure, attributes or items, levels, and response format are preserved. Line breaks, spacing, font size, and other visual layout elements may differ from the original Google Forms display. **Abbreviations**: AD, Alzheimer disease; ARIA, amyloid-related imaging abnormalities; BWS, best-worst scaling; DCE, discrete choice experiment; SDM, shared decision-making.

**Table 1.**
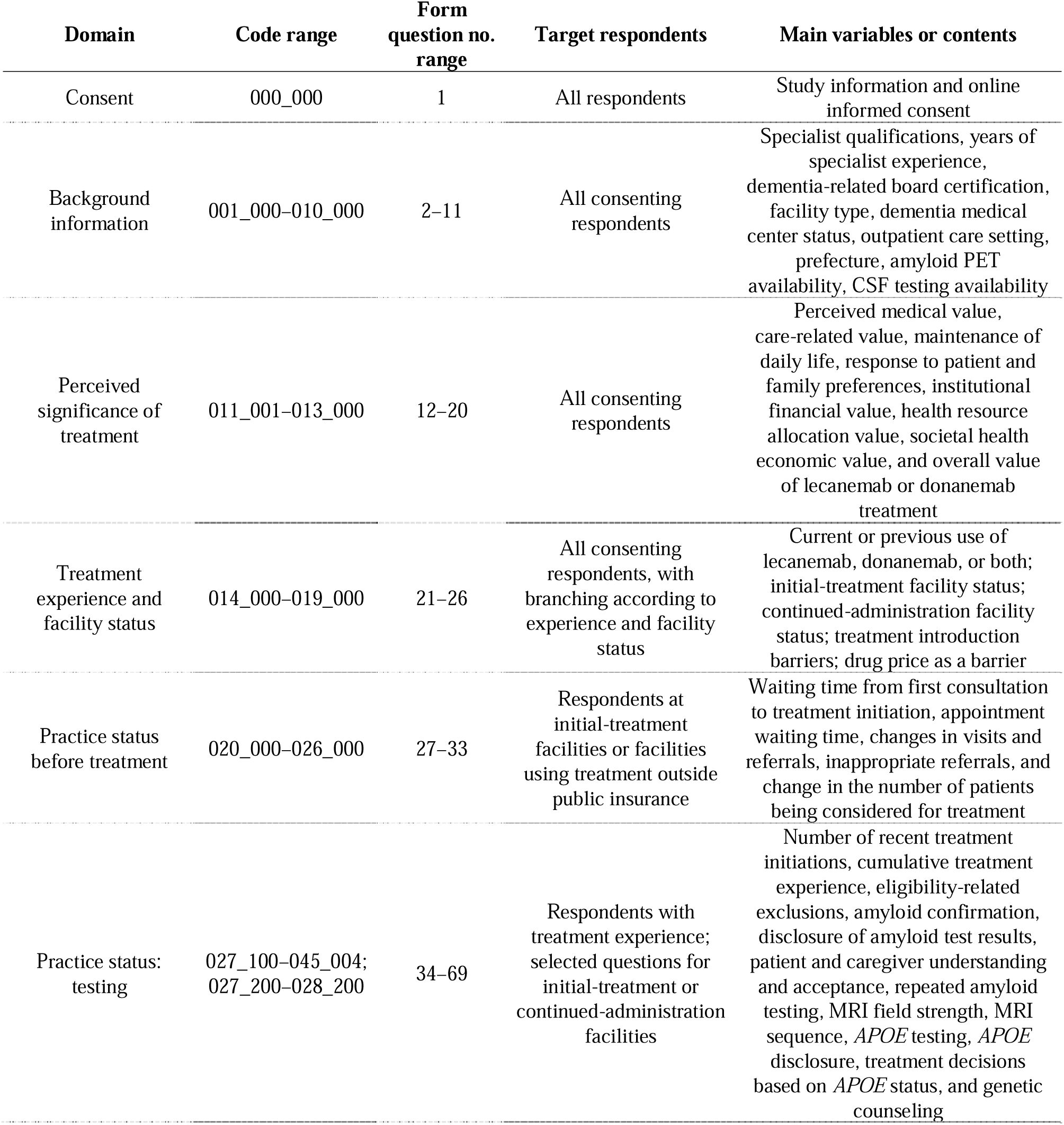
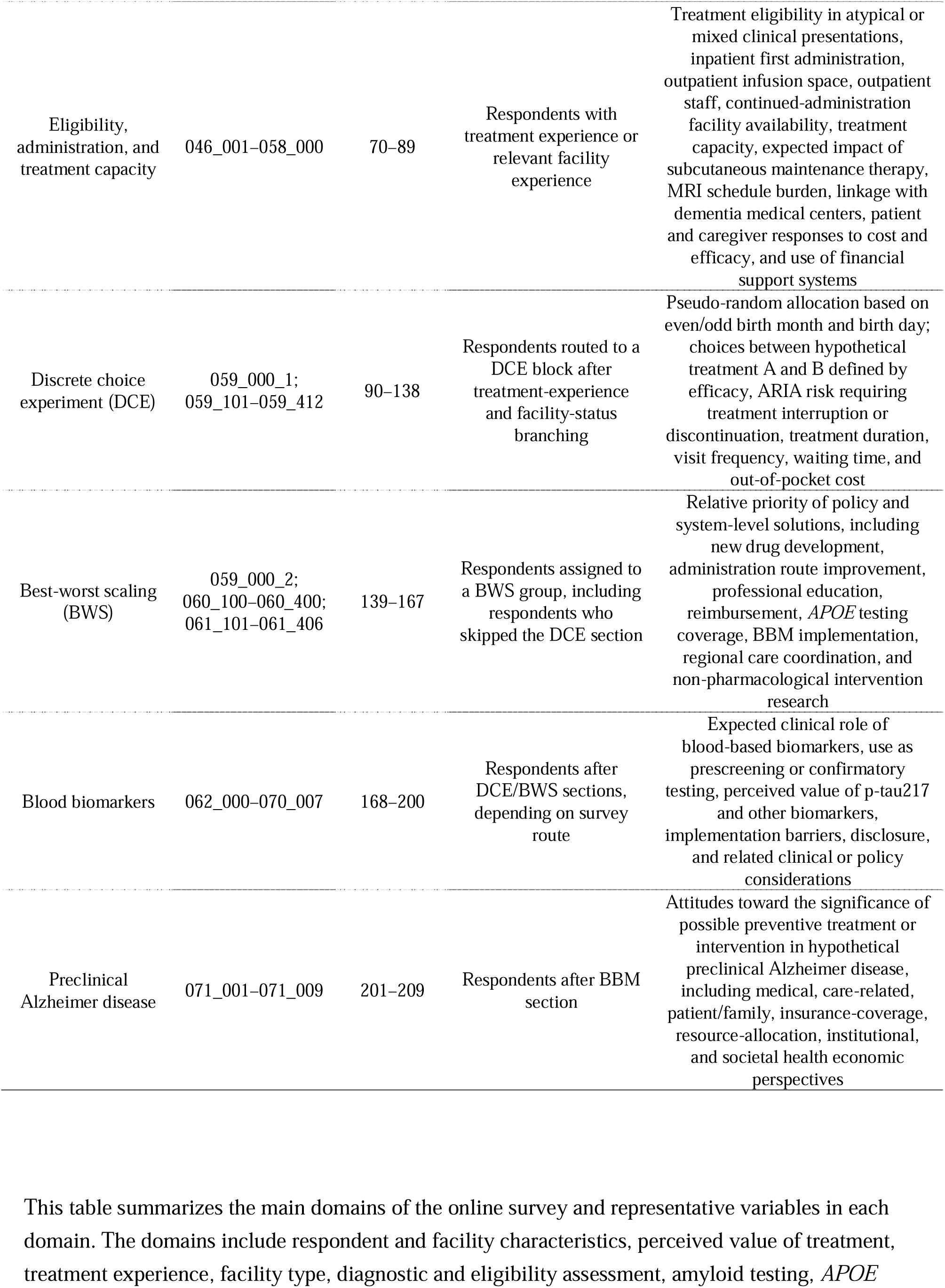
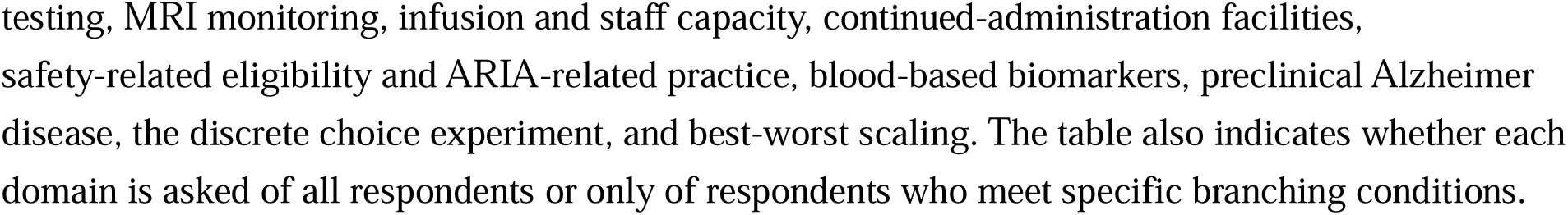
Survey domains and planned variables.

| Domain | Code range | Form question no. range | Target respondents | Main variables or contents |
| --- | --- | --- | --- | --- |
| Consent | 000_000 | 1 | All respondents | Study information and online informed consent |
| Background information | 001_000–010_000 | 2–11 | All consenting respondents | Specialist qualifications, years of specialist experience, dementia-related board certification, facility type, dementia medical center status, outpatient care setting, prefecture, amyloid PET availability, CSF testing availability |
| Perceived significance of treatment | 011_001–013_000 | 12–20 | All consenting respondents | Perceived medical value, care-related value, maintenance of daily life, response to patient and family preferences, institutional financial value, health resource allocation value, societal health economic value, and overall value of lecanemab or donanemab treatment |
| Treatment experience and facility status | 014_000–019_000 | 21–26 | All consenting respondents, with branching according to experience and facility status | Current or previous use of lecanemab, donanemab, or both; initial-treatment facility status; continued-administration facility status; treatment introduction barriers; drug price as a barrier |
| Practice status before treatment | 020_000–026_000 | 27–33 | Respondents at initial-treatment facilities or facilities using treatment outside public insurance | Waiting time from first consultation to treatment initiation, appointment waiting time, changes in visits and referrals, inappropriate referrals, and change in the number of patients being considered for treatment |
| Practice status: testing | 027_100–045_004;<br>027_200–028_200 | 34–69 | Respondents with treatment experience; selected questions for initial-treatment or continued-administration facilities | Number of recent treatment initiations, cumulative treatment experience, eligibility-related exclusions, amyloid confirmation, disclosure of amyloid test results, patient and caregiver understanding and acceptance, repeated amyloid testing, MRI field strength, MRI sequence, APOE testing, APOE disclosure, treatment decisions based on APOE status, and genetic counseling |
| Eligibility, administration, and treatment capacity | 046_001–058_000 | 70–89 | Respondents with treatment experience or relevant facility experience | Treatment eligibility in atypical or mixed clinical presentations, inpatient first administration, outpatient infusion space, outpatient staff, continued-administration facility availability, treatment capacity, expected impact of subcutaneous maintenance therapy, MRI schedule burden, linkage with dementia medical centers, patient and caregiver responses to cost and efficacy, and use of financial support systems |
| Discrete choice experiment (DCE) | 059_000_1;<br>059_101–059_412 | 90–138 | Respondents routed to a DCE block after treatment-experience and facility-status branching | Pseudo-random allocation based on even/odd birth month and birth day; choices between hypothetical treatment A and B defined by efficacy, ARIA risk requiring treatment interruption or discontinuation, treatment duration, visit frequency, waiting time, and out-of-pocket cost |
| Best-worst scaling (BWS) | 059_000_2;<br>060_100–060_400;<br>061_101–061_406 | 139–167 | Respondents assigned to a BWS group, including respondents who skipped the DCE section | Relative priority of policy and system-level solutions, including new drug development, administration route improvement, professional education, reimbursement, APOE testing coverage, BBM implementation, regional care coordination, and non-pharmacological intervention research |
| Blood biomarkers | 062_000–070_007 | 168–200 | Respondents after DCE/BWS sections, depending on survey route | Expected clinical role of blood-based biomarkers, use as prescreening or confirmatory testing, perceived value of p-tau217 and other biomarkers, implementation barriers, disclosure, and related clinical or policy considerations |
| Preclinical Alzheimer disease | 071_001–071_009 | 201–209 | Respondents after BBM section | Attitudes toward the significance of possible preventive treatment or intervention in hypothetical preclinical Alzheimer disease, including medical, care-related, patient/family, insurance-coverage, resource-allocation, institutional, and societal health economic perspectives |

This table summarizes the main domains of the online survey and representative variables in each domain. The domains include respondent and facility characteristics, perceived value of treatment, treatment experience, facility type, diagnostic and eligibility assessment, amyloid testing, APOE
testing, MRI monitoring, infusion and staff capacity, continued-administration facilities, safety-related eligibility and ARIA-related practice, blood-based biomarkers, preclinical Alzheimer disease, the discrete choice experiment, and best-worst scaling. The table also indicates whether each domain is asked of all respondents or only of respondents who meet specific branching conditions.

**Table 2.**
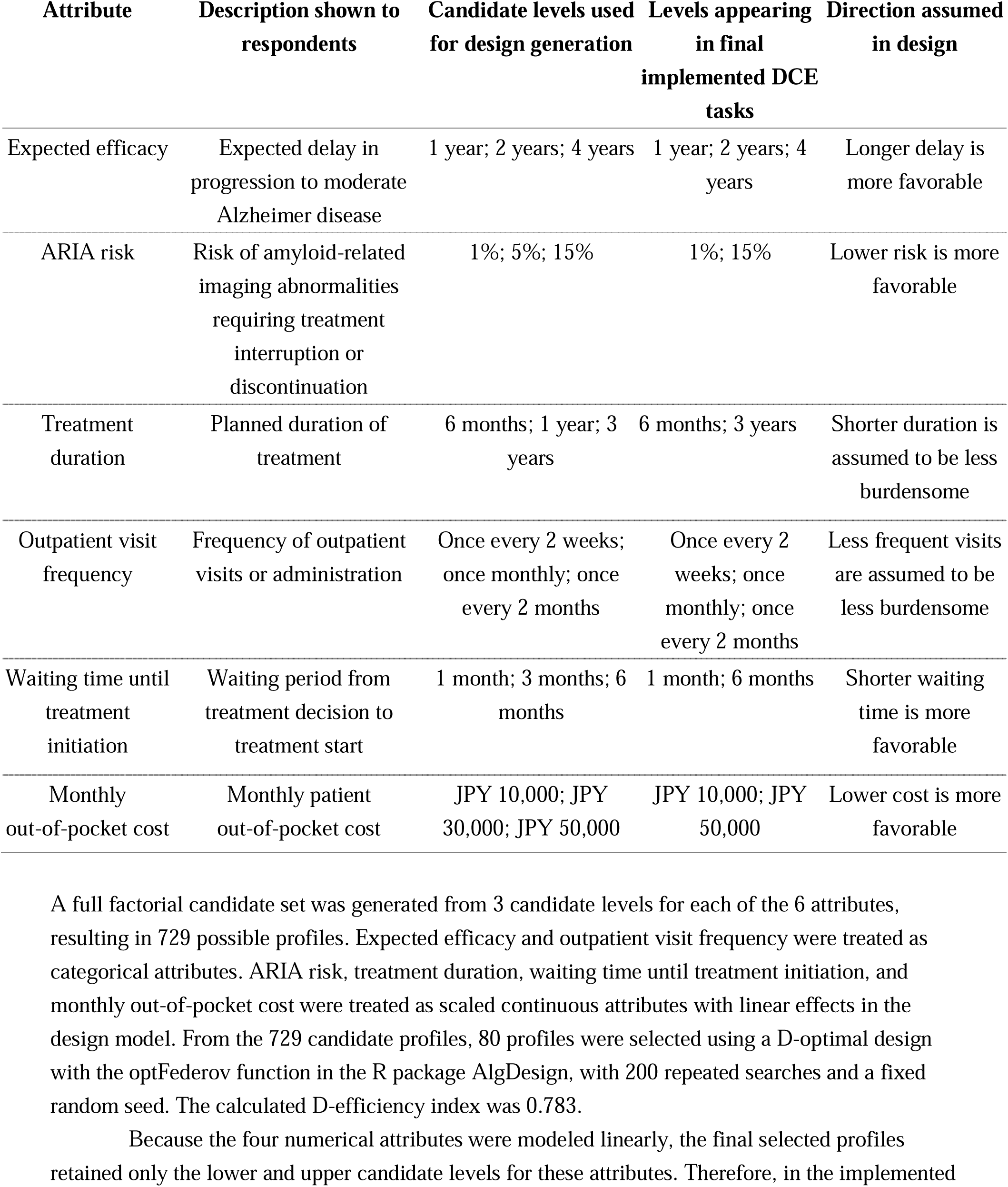
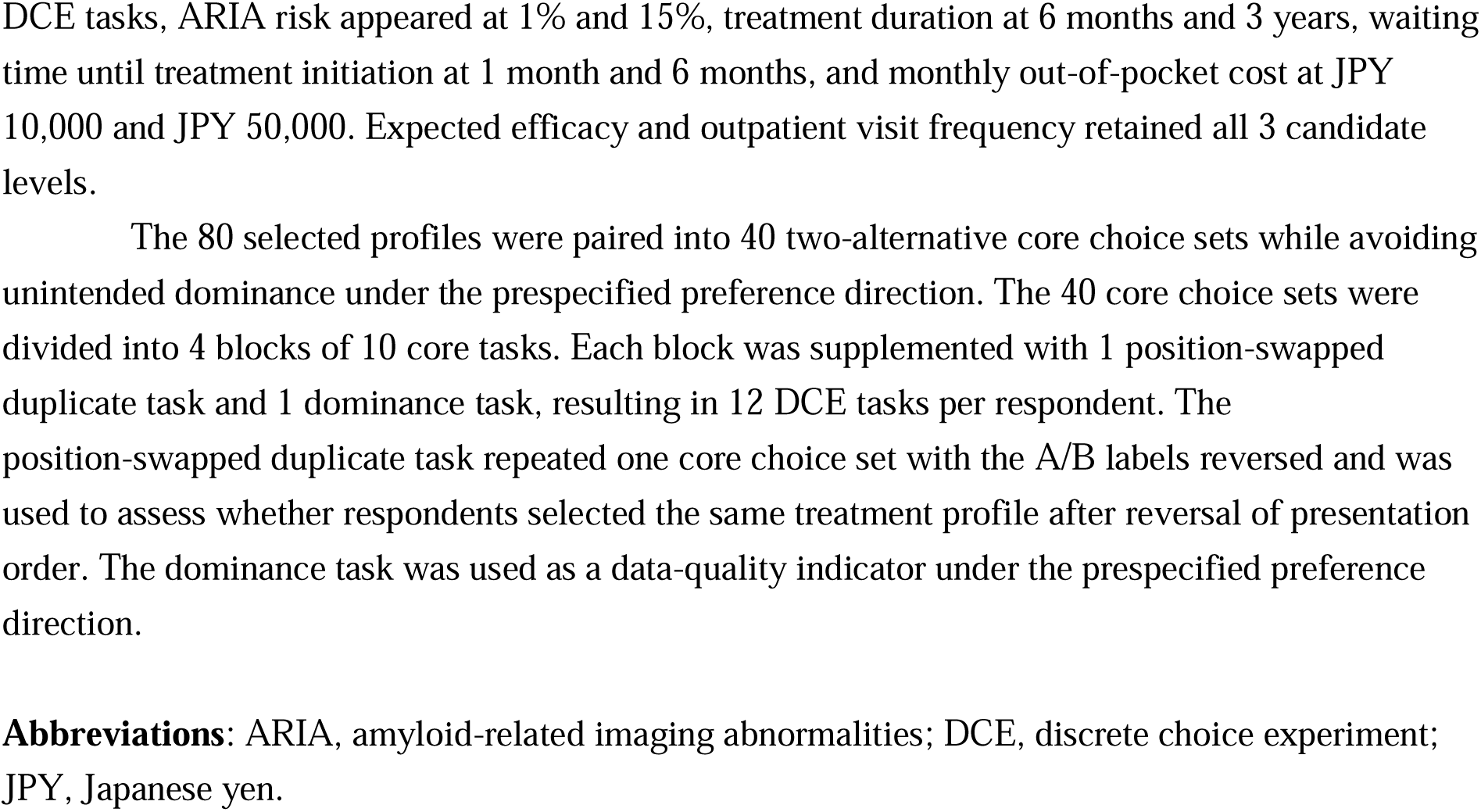
Attributes, candidate levels, and implemented levels used in the discrete choice experiment.

| Attribute | Description shown to respondents | Candidate levels used for design generation | Levels appearing in final implemented DCE tasks | Direction assumed in design |
| --- | --- | --- | --- | --- |
| Expected efficacy | Expected delay in progression to moderate Alzheimer disease | 1 year; 2 years; 4 years | 1 year; 2 years; 4 years | Longer delay is more favorable |
| ARIA risk | Risk of amyloid-related imaging abnormalities requiring treatment interruption or discontinuation | 1%; 5%; 15% | 1%; 15% | Lower risk is more favorable |
| Treatment duration | Planned duration of treatment | 6 months; 1 year; 3 years | 6 months; 3 years | Shorter duration is assumed to be less burdensome |
| Outpatient visit frequency | Frequency of outpatient visits or administration | Once every 2 weeks; once monthly; once every 2 months | Once every 2 weeks; once monthly; once every 2 months | Less frequent visits are assumed to be less burdensome |
| Waiting time until treatment initiation | Waiting period from treatment decision to treatment start | 1 month; 3 months; 6 months | 1 month; 6 months | Shorter waiting time is more favorable |
| Monthly out-of-pocket cost | Monthly patient out-of-pocket cost | JPY 10,000; JPY 30,000; JPY 50,000 | JPY 10,000; JPY 50,000 | Lower cost is more favorable |

A full factorial candidate set was generated from 3 candidate levels for each of the 6 attributes, resulting in 729 possible profiles. Expected efficacy and outpatient visit frequency were treated as categorical attributes. ARIA risk, treatment duration, waiting time until treatment initiation, and monthly out-of-pocket cost were treated as scaled continuous attributes with linear effects in the design model. From the 729 candidate profiles, 80 profiles were selected using a D-optimal design with the optFederov function in the R package AlgDesign, with 200 repeated searches and a fixed random seed. The calculated D-efficiency index was 0.783.
Because the four numerical attributes were modeled linearly, the final selected profiles retained only the lower and upper candidate levels for these attributes. Therefore, in the implemented
DCE tasks, ARIA risk appeared at 1% and 15%, treatment duration at 6 months and 3 years, waiting time until treatment initiation at 1 month and 6 months, and monthly out-of-pocket cost at JPY 10,000 and JPY 50,000. Expected efficacy and outpatient visit frequency retained all 3 candidate levels.
The 80 selected profiles were paired into 40 two-alternative core choice sets while avoiding unintended dominance under the prespecified preference direction. The 40 core choice sets were divided into 4 blocks of 10 core tasks. Each block was supplemented with 1 position-swapped duplicate task and 1 dominance task, resulting in 12 DCE tasks per respondent. The position-swapped duplicate task repeated one core choice set with the A/B labels reversed and was used to assess whether respondents selected the same treatment profile after reversal of presentation order. The dominance task was used as a data-quality indicator under the prespecified preference direction.
**Abbreviations:** ARIA, amyloid-related imaging abnormalities; DCE, discrete choice experiment; JPY, Japanese yen.

**Table 3.**
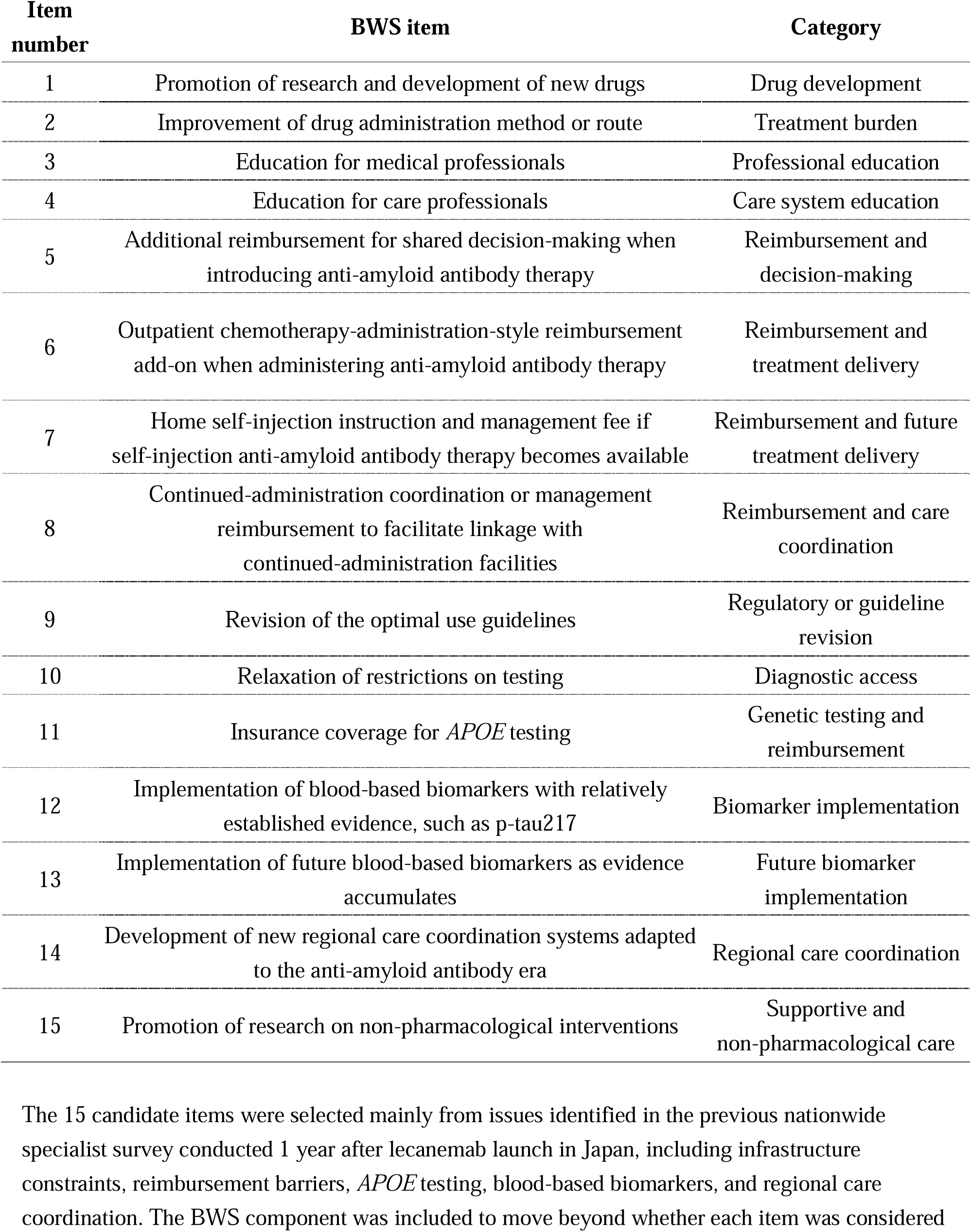
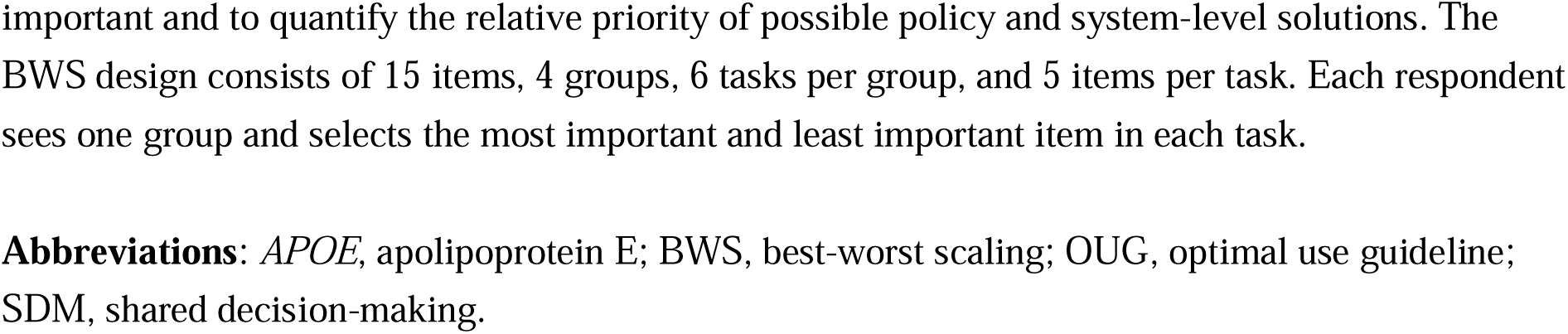
Items included in the best-worst scaling component.

| Item number | BWS item | Category |
| --- | --- | --- |
| 1 | Promotion of research and development of new drugs | Drug development |
| 2 | Improvement of drug administration method or route | Treatment burden |
| 3 | Education for medical professionals | Professional education |
| 4 | Education for care professionals | Care system education |
| 5 | Additional reimbursement for shared decision-making when introducing anti-amyloid antibody therapy | Reimbursement and decision-making |
| 6 | Outpatient chemotherapy-administration-style reimbursement add-on when administering anti-amyloid antibody therapy | Reimbursement and treatment delivery |
| 7 | Home self-injection instruction and management fee if self-injection anti-amyloid antibody therapy becomes available | Reimbursement and future treatment delivery |
| 8 | Continued-administration coordination or management reimbursement to facilitate linkage with continued-administration facilities | Reimbursement and care coordination |
| 9 | Revision of the optimal use guidelines | Regulatory or guideline revision |
| 10 | Relaxation of restrictions on testing | Diagnostic access |
| 11 | Insurance coverage for *APOE* testing | Genetic testing and reimbursement |
| 12 | Implementation of blood-based biomarkers with relatively established evidence, such as p-tau217 | Biomarker implementation |
| 13 | Implementation of future blood-based biomarkers as evidence accumulates | Future biomarker implementation |
| 14 | Development of new regional care coordination systems adapted to the anti-amyloid antibody era | Regional care coordination |
| 15 | Promotion of research on non-pharmacological interventions | Supportive and non-pharmacological care |

The 15 candidate items were selected mainly from issues identified in the previous nationwide specialist survey conducted 1 year after lecanemab launch in Japan, including infrastructure constraints, reimbursement barriers, *APOE* testing, blood-based biomarkers, and regional care coordination. The BWS component was included to move beyond whether each item was considered
important and to quantify the relative priority of possible policy and system-level solutions. The BWS design consists of 15 items, 4 groups, 6 tasks per group, and 5 items per task. Each respondent sees one group and selects the most important and least important item in each task.
**Abbreviations:** *APOE*, apolipoprotein E; BWS, best-worst scaling; OUG, optimal use guideline; SDM, shared decision-making.

**Table 4.** Planned outcomes and analysis approach.

| Outcome category | Planned outcomes | Main variables | Planned analysis |
| --- | --- | --- | --- |
| Primary outcomes | Current implementation status of anti-amyloid antibody therapy | Current or previous use of lecanemab, donanemab, or both; facility ability to provide initial treatment or continued administration | Descriptive statistics; stratified analysis by treatment experience and facility type |
| Primary outcomes | Facility readiness and implementation barriers | Amyloid PET availability, CSF testing availability, MRI monitoring, ARIA response capacity, outpatient infusion space, staff resources, continued-administration facility availability | Counts and percentages; stratified analysis; exploratory logistic regression when appropriate |
| Primary outcomes | Diagnostic and treatment workflow | Waiting time from first consultation to treatment initiation, appointment waiting time, number of recent treatment initiations, patient flow, referral patterns | Descriptive statistics using median and interquartile range or counts and percentages |
| Primary outcomes | Support for policy and system-level solutions | Support for reimbursement, *APOE* testing coverage, BBM implementation, regional care coordination, OUG revision, education, and other proposed solutions | Counts and percentages; exploratory regression models for selected outcomes |
| Secondary outcomes | Treatment preference in the DCE | Choices between hypothetical treatment A and B; attributes including efficacy, ARIA risk requiring treatment interruption or discontinuation, treatment duration, visit frequency, waiting time, and out-of-pocket cost | Conditional logit model as primary approach; mixed logit model if sample size is sufficient; sensitivity analysis using data quality indicators |
| Secondary outcomes | Relative priority of policy solutions in BWS | Best and worst selections among 15 policy and system-level items | Count-based best-minus-worst scores; conditional logit model when appropriate; ranking of items |
| Secondary outcomes | Attitudes toward *APOE* testing | Current use of *APOE* testing, disclosure of results, genetic counseling, treatment decisions based on *APOE* status, perceived clinical value | Descriptive statistics; stratified analysis by treatment experience and facility type |
| Secondary outcomes | Attitudes toward blood-based biomarkers | Expected use as prescreening, confirmatory testing, or future implementation tool; perceived value of p-tau217 and other biomarkers | Descriptive statistics; exploratory group comparisons |
| Secondary outcomes | Attitudes toward preclinical Alzheimer disease | Views on the significance of possible preventive treatment or intervention in hypothetical preclinical Alzheimer disease from medical, care-related, patient/family, insurance-coverage, resource-allocation, institutional, and societal health economic perspectives | Descriptive statistics; exploratory group comparisons |
| Feasibility outcomes | Survey feasibility and completeness | Number of responses, completion rate, item-level missingness, section-level missingness, branching-related missingness | Descriptive statistics; missingness summary by domain |
| Feasibility outcomes | DCE and BWS feasibility | DCE response rate, BWS response rate, time or completion status if available | Descriptive statistics |
| Data quality outcomes | Internal consistency and attention checks | Dominance task result, position-swapped duplicate task consistency, and instructed-response attention-check item | Proportion passing the dominance task; proportion selecting the same treatment profile in position-swapped duplicate tasks; proportion passing the instructed-response attention-check item; sensitivity analyses excluding low-quality responses |
| Missing data handling | Handling of incomplete or non-applicable responses | Structural missingness due to branching, optional item nonresponse, invalid or uninterpretable responses | Primary analysis using available valid responses for each item; complete-case analysis for regression models; sensitivity analyses if needed |

This is an exploratory survey protocol. The primary analyses will be descriptive. Regression, DCE, and BWS analyses will be interpreted as exploratory. Percentages will generally use the number of valid responses to each question as the denominator, with structural missingness due to branching distinguished from optional nonresponse.
**Abbreviations:** APOE, apolipoprotein E; ARIA, amyloid-related imaging abnormalities; BBM, blood-based biomarker; BWS, best-worst scaling; CSF, cerebrospinal fluid; DCE, discrete choice experiment; MRI, magnetic resonance imaging; OUG, optimal use guideline; PET, positron emission tomography.

## Acknowledgements

This survey was carried out by the FY2025-2026 Ministry of Health, Labour and Welfare (MHLW) Scientific Research Project titled “*A Study on Responses to Social Issues Arising from Advances in Dementia Medicine*” (principal investigator (PI): Tetsuaki Arai), in collaboration with the Japanese Society for Dementia Research (president: Takeshi Ikeuchi) and the Japanese Psychogeriatric Society (president: Manabu Ikeda).

Some authors’ affiliation (KS, YN, TI), “*Dementia Inclusion and Therapeutics*,” is an endowed department at the University of Tokyo Hospital funded by Effissimo Capital Management Pte Ltd.

AI-assisted tools, including ChatGPT and Claude, were used to support English language editing and to check clarity and consistency of the manuscript. These tools were not used to generate or analyze survey data. All authors reviewed and approved the final manuscript, and the authors take full responsibility for the content.

## Consent Statement

Online informed consent was obtained on the introductory page of the survey prior to questions.

## Data Availability

The deidentified survey dataset generated in this study will not be publicly shared. Aggregated results will be reported in the final study publication.

