## Supplementary table Legend for "Protocol for the 2026 Nationwide Survey of Dementia Specialists on the Real-World Implementation of Anti-Amyloid Antibody Therapies in Japan: Clinical Practice, Blood Biomarkers, and Preference Experiments"

Supplementary Materials

**Table S1. Detailed questionnaire in Japanese and English**

Table S1 contains the full questionnaire, including the original Japanese text, English translation, question codes, response formats, response options, mandatory status, branching logic, and interpretation notes. Note: The BWS allocation question is logically used before the BWS tasks to route respondents to one of four BWS presentation groups. Its form question number reflects its physical position in the exported Google Forms structure.

**Table S2. Master table for the discrete choice experiment**

Table S2 lists the DCE attributes, candidate levels, implemented levels, analysis coding, and prespecified preference direction.

**Table S3. Design matrix for the discrete choice experiment**

Table S3 shows the full DCE design matrix, including block number, task type, duplicate-task linkage, dominance-task status, alternative labels, and attribute levels.

**Table S4. Master table for the best-worst scaling component**

Table S4 lists the 15 BWS items in Japanese and English, together with item categories and notes on item selection.

**Table S5. Design matrix for the best-worst scaling component**

Table S5 shows the BWS design matrix, including group, task, item order, item number, item labels, and best/worst response variable names.
